# The Citation Cloud of a Biomedical Article: Enabling Citation Analysis

**DOI:** 10.1101/2020.06.15.20131623

**Authors:** Neil R. Smalheiser, Jodi Schneider, Vetle I. Torvik, Dean P. Fragnito, Eric E. Tirk

## Abstract

Using open citations provided by iCite and other sources, we have built an extension to PubMed that allows any user to visualize and analyze the “citation cloud” around any target article A: the set of articles cited by A; those which cite A; those which are co-cited with A; and those which are bibliographically coupled to A. This greatly enables the study of citations by the scientific community. The Citation Cloud can be accessed by running any query on the Anne O’Tate value-added PubMed search interface http://arrowsmith.psych.uic.edu/cgi-bin/arrowsmith_uic/AnneOTate.cgi and clicking on the Citations button next to any retrieved article.

## Introduction

Citation analysis is crucial for tracing the diffusion of knowledge across disciplines and over time, both at the micro and macro level. For example, one may wish to follow citation chains, e.g., identifying the influence of a retracted article on later papers that cite it [1,2]. Hutchins et al have employed citation patterns to predict which articles are likely to contribute to translation of basic studies into clinical advances [3]. More globally, Boyack and Klavans employed citations to identify research frontiers [4].

Citation analysis has largely been the province of scholars in the specialties of bibliometrics, scientometrics, innovation and policy studies, who typically carry out extensive manual analysis of proprietary citation data licensed by commercial data providers. This has limited the extent to which the scientific community can utilize citations. Recently, iCite, an extensive set of open citations in the biomedical literature, has been publicly released [5] and the dataset is updated monthly (https://icite.od.nih.gov/). This provides a great opportunity for biomedical investigators and other interested parties, but to date, there is no user-friendly interface for accessing or analyzing the citation data. Here, we describe Citation Cloud, an extension to PubMed (the leading public biomedical search engine https://www.ncbi.nlm.nih.gov/pubmed/) that allows any user to visualize and analyze the “citation cloud” around any target article A: the set of articles cited by A; those which cite A; those which are co-cited with A; and those which are bibliographically coupled to A.

To say that an article B is co-cited with the target article A means that they are both cited by the same article(s) Ci [6]. Co-citation is a measure of similarity not based on textual or topical similarity. Note that the co-citation relationship is not fixed but can vary over time depending on how many newer articles cite both A and B. In contrast, bibliographically coupled articles cite some of the same articles in their reference lists as does the target article A [7]. In other words, their reference lists overlap. This is also a measure of similarity that is not based on textual or topical similarity, and has the distinct advantage that the bibliographically coupled relationship can be calculated for any two articles regardless of when they are published. As well, this relationship is stable and will not change over time.

### How Citation Cloud works

The Citation Cloud can be accessed by running any query on the Anne O’Tate value-added PubMed search interface http://arrowsmith.psych.uic.edu/cgi-bin/arrowsmith_uic/AnneOTate.cgi <u>[8</u>, ms. submitted] and clicking on the Citations button next to any retrieved article. For example, suppose we enter the query “Retractions in the medical literature: how many patients are put at risk by flawed research?” to retrieve this single article (Fig. 1).

**Figure 1.**
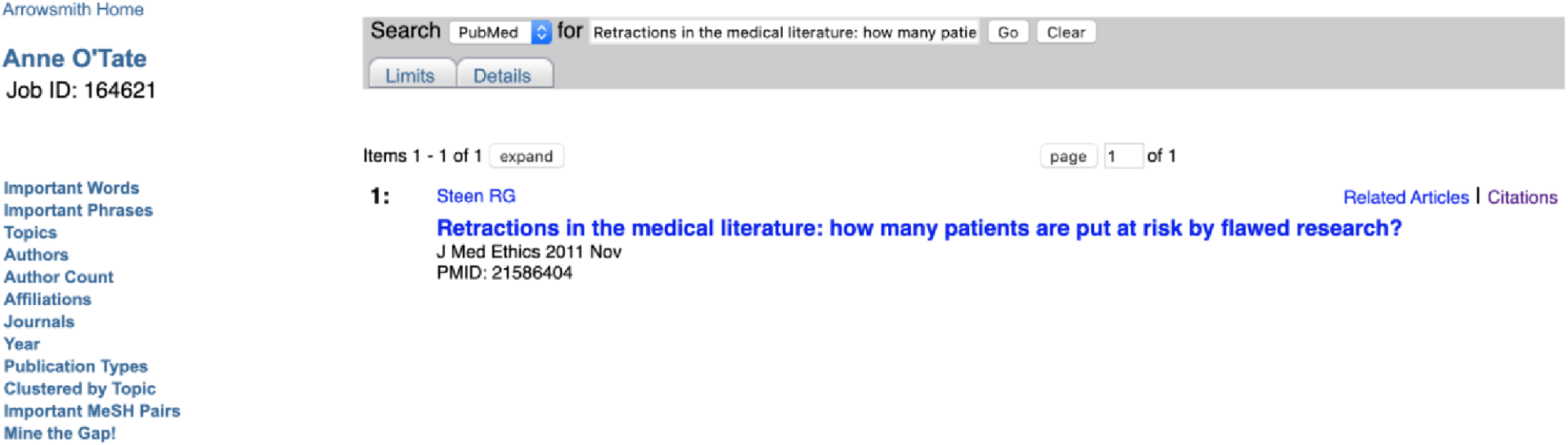
Screenshot of a PubMed query entered via the Anne O’Tate tool.

Shown is the article retrieved by using the title in the query box. The hotlinked word Citations is displayed to the right of the article.

Click on the Citations button next to it and we see the following screenshot on a new tab (Fig. 2).

**Figure 2.**
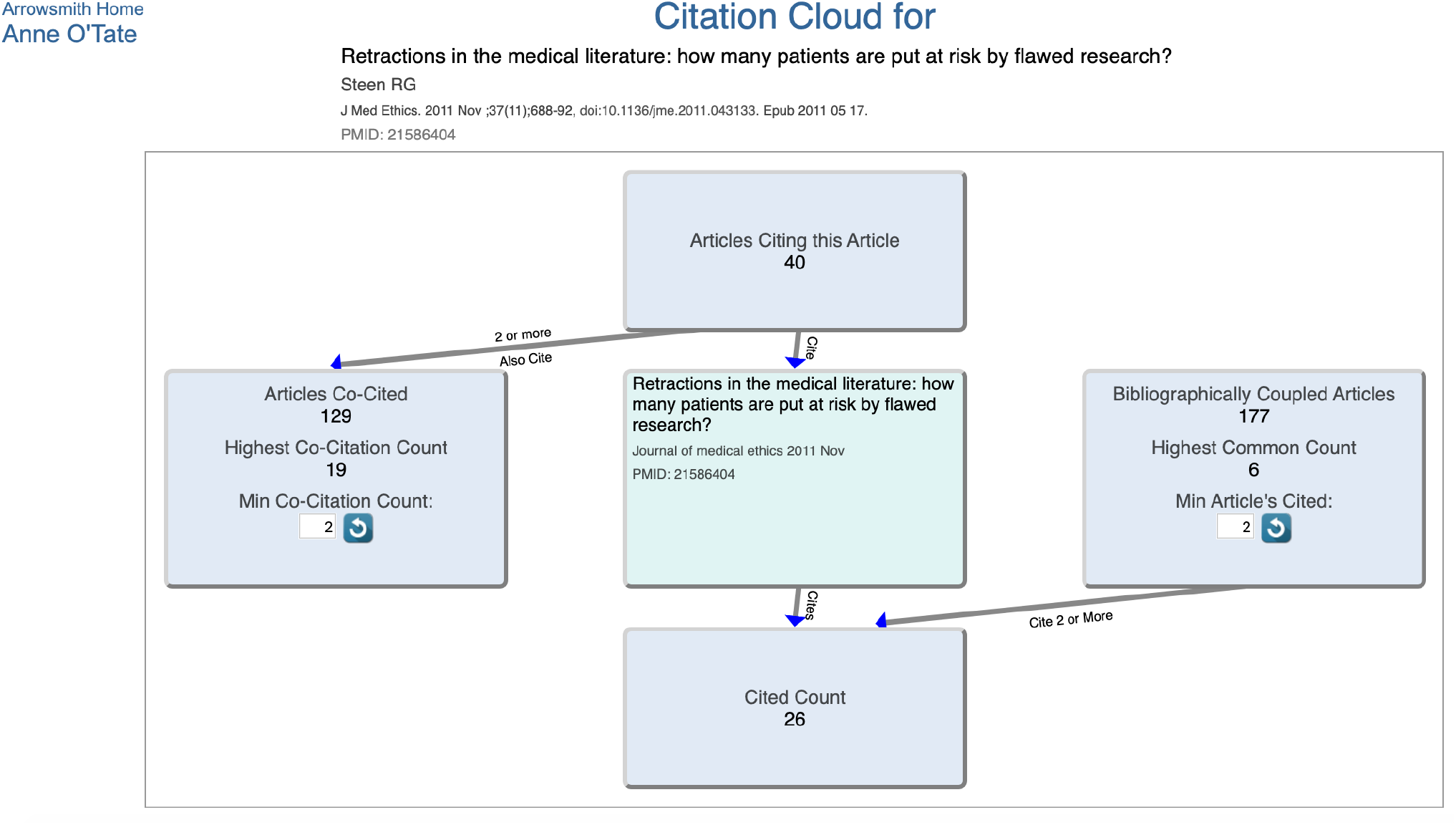
The Citation Cloud visualization for the article displayed in Figure 1.

The Citation Cloud consists of five boxes that are interlinked by arrows that show the direction of citations. Clicking on any box opens a new tab that shows the articles in that box, and has hotlinks to allow users to export the articles to PubMed or Anne O’Tate for further mining.

The target article is in the center box.

The “Articles Citing this Article” box consists of all articles that cite the target article. In this example, there are 40 citing articles.

The “Cited Count” box consists of all articles in the reference list of the target article.

The “Articles Co-Cited” box consists of all articles that are cited by one or more papers in the “Articles Citing this Article” box. The default co-citation count threshold for displaying articles in this box is 2 – that means that each article displayed in the “Articles Co-Cited” box is cited by at least 2 articles in the “Article Citing this Article” box. Highly cited target articles may have a very large set of co-cited articles, so we allow users to adjust the co-citation count threshold as desired.

The “Bibliographically Coupled Articles” box consists of all articles that cite papers in the reference list of the target article. The default threshold for displaying articles in this box is 2 – that means that each article displayed in the “Bibliographically Coupled Articles” box cites at least 2 different articles in the reference list of the target article. Again, users can adjust the threshold.

The target article is in the center box, with four boxes surrounding it. The upper box shows that 40 articles have cited the target article; clicking on this box opens a new Results tab that lists the 40 articles (Fig. 3).

**Figure 3.**
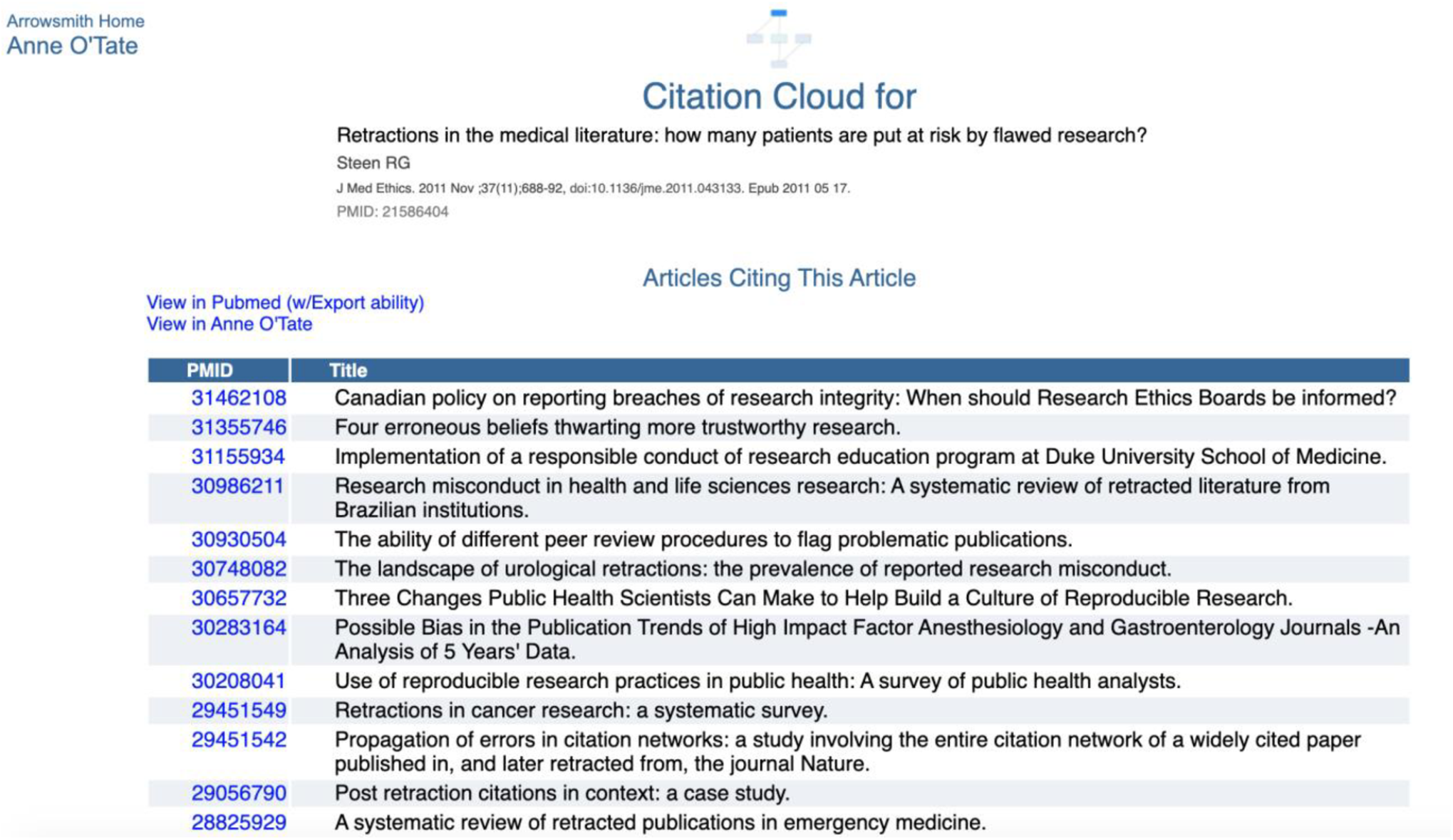
Screenshot of the contents of the “Articles Citing this Article” box.

Similarly, by clicking on the respective boxes, one can view and process articles that are cited by the target article; that are co-cited; or that are bibliographically coupled. The default option is to display a threshold of two – this means that at least two articles in the “citing” box cited any article displayed in the “co-cited” box; conversely, for the “bibliographically coupled” box that means that each displayed bibliographically coupled article cited at least two references within the “cited “box. The minimum threshold for display can be varied by the user, in order to focus on the articles having the most similarity to the target article. Each box has two hot links that permit the user to export the list to PubMed (which has the ability to export the citations in various formats) or to export the list to Anne O’Tate [8] where it can be mined further. For example, one can identify the most important words and phrases in the titles and abstracts of articles on the list, as well as the most frequent topics, authors, journals, etc. [8].

### Limitations

The initial dataset of open citations, while reasonably up to date, is static. The iCite dataset is updated monthly and these new citations will be automatically added to our dataset. However, since not all citations are openly available [12], the set of citations is far from comprehensive. Whereas we incorporated citations from over 17 million unique articles indexed in PubMed, including proprietary citations from Web of Science and Scopus would have given access to ∼21 million articles. Another limitation is that the citation cloud surrounding a single article can be quite large, especially for review articles or citation classics. Thus, it may be too cumbersome to display a citation cloud to encompass an entire list of articles.

### Benefits to the Community

We expect that this new tool will augment the power of the new open citations datasets to enable a broad community of scientists to utilize citations in their studies of biomedical literature.

The citation cloud may be useful to biomedical investigators and public users who are not carrying out citation analysis per se. The co-cited and bibliographically coupled articles represent types of similarity that are complementary to the PubMed Related Articles ranking [9], and thus may assist in increasing recall for information retrieval [10], for example, in finding relevant literature for writing systematic reviews [11].

## Materials and Methods

We seeded the Citation Cloud dataset with a very extensive set of open citations which was culled from six different sources (Table 1), and included the initial release of the NIH iCite dataset in October 2019 [5].

**Table 1.**
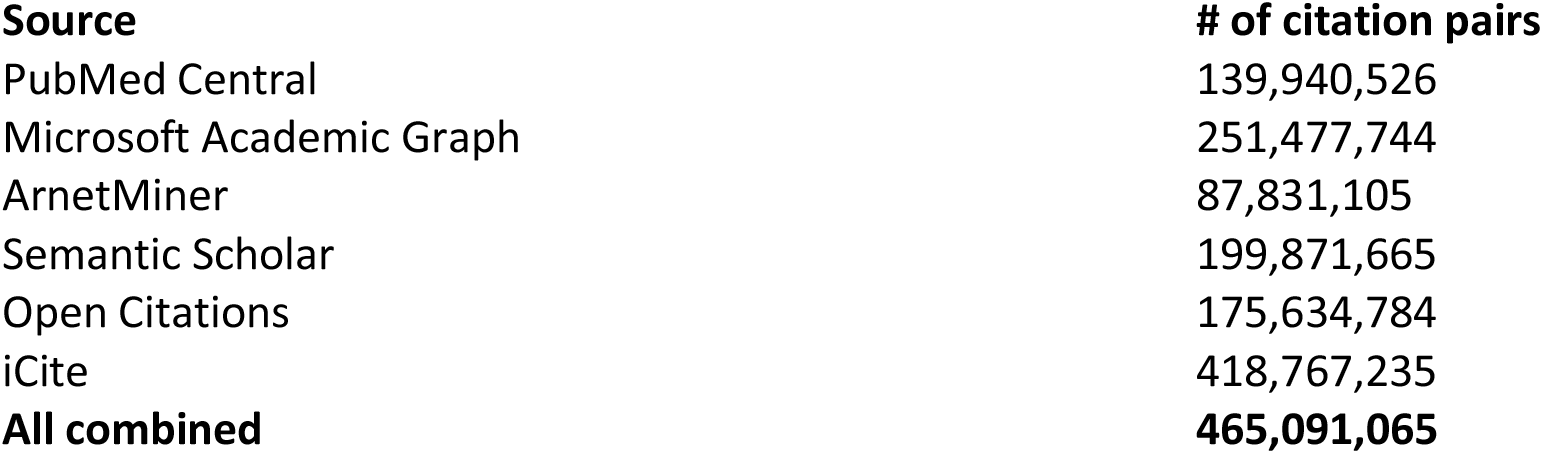
Initial dataset of open citations to seed the Citation Cloud.

| **Source** | **# of citation pairs** |
| --- | --- |
| PubMed Central | 139,940,526 |
| Microsoft Academic Graph | 251,477,744 |
| ArnetMiner | 87,831,105 |
| Semantic Scholar | 199,871,665 |
| Open Citations | 175,634,784 |
| iCite | 418,767,235 |
| **All combined** | **465,091,065** |

The combined set represents the union of citation pairs (article A cites article B) for all six sources after removing duplicates. The article pairs comprise 17,681,409 unique PMIDs.

### EAV database architecture

An Entity-Attribute-Value (EAV) database structure was created to enable the generic storing and efficient querying of the sparsely populated PubMed source XML documents. It stores both single and multi-valued elements, naturally handles sparse data (not all documents contain all possible elements), and stores new elements (previously not found in a PubMed source document) without the need for database modification such as table creation, column creation, or index creation.

Traditional relational database structures use the technique of normalizing multi-valued elements into separate tables. There are positives and negatives to this. On the positive side, indexes may be created on the columns belonging to a multi-valued element. On the negative side, the normalizing process requires work to design the tables and design the indexes and requires custom coding to extract and populate the tables. Additionally, with sparse datasets such (i.e. PubMed) normalized models suffer from inefficiencies in disk usage and thus query performance, due to many empty or NULL values where no values exist in the source data (e.g. no FirstName in source but having a dedicated FirstName column in the database).

The EAV model as implemented combines the positives of indexing and none of the design and extract work. However it does require that relatively more complex SQL queries be written.

Some EAV systems store all data - regardless of type (string, number, date, etc) - as a string. This leads to problems when for instance, one is trying to sort by a number field but the values are stored as strings, in which case the query would sort incorrectly (e.g. 1, 12, 2, 200, 3…).

Other EAV systems store different data types in specific columns that match the incoming data type. This is more efficient in terms of index use and query speed, but makes for even more complex queries and updates. Our EAV database structure incorporates a novel technique against two specific data types (strings and integers) that allows for simple storage to a single string column, yet makes use of indexes specific to those two data types. Future versions can be expanded to handle other data types such as Dates.

The technique automatically stored strings into a virtual column named ‘valshort’ which is indexed to the first 40 characters of the original string value. This allows one to quickly search against string values known to be short (e.g. LastName) by querying against the database’s fully indexed valshort column rather than against the non-indexable large Text column ‘val’. Integer values are likewise automatically converted by the database from the original XML string values and stored and indexed to the virtual ‘valint’ column. These virtual columns do not cause redundant storage space to be consumed as would real columns. They only consume storage within their given indexes.

Here is an example of a query which returns the list of co-citers within the Citation Cloud:

~~~
SELECT t1.aid as ‘pmid’, citer4.val as ‘title’
FROM (
  SELECT
  citer.aid as aid
  FROM aelement as citer
  JOIN eir en2 ON
    en2.hs=‘/PubmedArticle/PubmedData/ReferenceList/Reference/ArticleIdList/ArticleId’
  WHERE citer.eirid=en2.eirid AND citer.valint=20072710
) as t1
LEFT JOIN eir en4 ON en4.hs=‘/PubmedArticle/MedlineCitation/Article/ArticleTitle’
LEFT JOIN aelement as citer4 ON citer4.aid=t1.aid and citer4.eirid=en4.eirid
ORDER BY pmid DESC
~~~

which returns (1st 10 shown):

~~~
+--------------+----------------------------------------------------------------------------------------------------
--+
|     pmid | title                                 |
+--------------+----------------------------------------------------------------------------------------------------
--+
|    30256792 | Self-citation is the hallmark of productive authors, of any gender.
|
|    30197432 | Last Place? The Intersection of Ethnicity, Gender, and Race in
Biomedical.           |
|    28771391 | Gender Differences in Receipt of National Institutes of Health R01 Grants Among Junior Faculty at… |
|    28758138 | Author Name Disambiguation for PubMed.
|
|    28509897 | Disambiguation of patent inventors and assignees using high-resolution
geolocation data.         |
|    28412964 | MeSH Now: automatic MeSH indexing at PubMed scale via learning to
rank.          |
|    27942200 | Quantifying Conceptual Novelty in the Biomedical Literature.
|
|    27457939 | Kin of coauthorship in five decades of health science literature.
|
|    27367860 | Author Disambiguation in PubMed: Evidence on the Precision and Recall
of Author-ity among NIH-Funded |
|    27213780 | Two Similarity Metrics for Medical Subject Headings (MeSH): An Aid to
Biomedical Text Mining and… |
…
~~~

Another example from the Citation Cloud returns a list of articles that are Bibliographically Coupled to pmid 20072710. The columns returned are pmid, title and common_article_count (the number of articles cited by the given pmid that are in common with the articles cited by the target pmid 20072710):

~~~
SELECT t1.aid_bca as ‘pmid’, caTitleEle.val as ‘title’, t1.bca_cac ‘common_article_count’
/* common article count */
FROM
(
       SELECT ca.aid as ‘aid_bca’, COUNT(ca.valint) as ‘bca_cac’ /* common article
count */
       FROM aelement PARTITION (active) as target
       JOIN eir en2 ON
en2.hs=‘/PubmedArticle/PubmedData/ReferenceList/Reference/ArticleIdList/ArticleId’
       /* join to other articles referencing same article as the target does */
       JOIN aelement ca ON ca.eirid=en2.eirid AND ca.valint=target.valint AND
ca.aid<>target.aid
       WHERE target.eirid=en2.eirid AND target.aid=20072710
       GROUP BY aid_bca
       HAVING bca_cac>=“.$MIN_bibc_count.”
       ORDER BY bca_cac DESC, aid_bca DESC
)
AS t1
/* join to title element of coupled article */
JOIN eir enTitle ON enTitle.hs=‘/PubmedArticle/MedlineCitation/Article/ArticleTitle’
LEFT JOIN aelement as caTitleEle ON caTitleEle.aid=aid_bca and
caTitleEle.eirid=enTitle.eirid
~~~

which returns (1st 10 shown):

~~~
+----------+--------------------------------------------------------------------------------------------------------
------+----------------------+
| pmid | title |
common_article_count |
+----------+--------------------------------------------------------------------------------------------------------
-----------------------------+
| 29271976 | Gaps within the Biomedical Literature: Initial Characterization and Assessment of Strategies for Discovery. |          3 |
| 25661592 | Context-driven automatic subgraph creation for literature-based discovery.
|          2 |
| 25472905 | Mammalian Argonaute-DNA binding?
|          2 |
| 24376375 | Studying PubMed usages in the field for complex problem solving:
Implications for tool design.          |          2 |
| 23894639 | Has large-scale named-entity network analysis been resting on a flawed
assumption?           |          2 |
| 22195132 | SEACOIN--an investigative tool for biomedical informatics researchers.
|          2 |
| 30533534 | Demystifying probabilistic linkage: Common myths and misconceptions.
|          1 |
| 30294517 | Knowledge-based biomedical Data Science.
|          1 |
| 30272675 | How user intelligence is improving PubMed.
|          1 |
| 30266789 | Literature-based automated discovery of tumor suppressor p53 phosphorylation and inhibition by NEK2.          |          1 |
…
~~~

In a non-EAV db configuration, we would have to create a secondary table to handle the many-to-many relationship sourced from PubMed’s /PubmedArticle/PubmedData/ReferenceList/Reference/ArticleIdList/ArticleId element, re-parse the entire corpus of PubMed just to get that one element, write dedicated code to deal with importing that element, and still have to JOIN to another table to get the Article Title to produce the above query results. With the EAV database structure, we had already imported every element from PubMed in a single generic routine so we already had the above element stored. No special tables or import routines were needed to deal with this (or any of the other multi-valued elements), and indexes were already created and optimized. Given these benefits, we anticipate greatly accelerated development time and little if any new database design and maintenance for future enhancements and projects.

The Architecture supporting The Citation Cloud is comprised of LINUX Ubuntu Server 18.04 LTS, Perl 5 version 26, and MySql 5.7.

## Data Availability

The Citation Cloud can be accessed by running any query on the Anne OTate value-added PubMed search interface http://arrowsmith.psych.uic.edu/cgi-bin/arrowsmith_uic/AnneOTate.cgi and clicking on the Citations button next to any retrieved article. The tool is free and public and requires no registration or password.

## Funding

This research was supported by NIH Grants R01LM010817 and P01AG039347. The study sponsor had no role in study design; in the collection, analysis and interpretation data; in the writing of the report; or in the decision to submit the paper for publication.

## Competing Interests

The authors declare no competing interests.

## Notes

### Competing Interest Statement

The authors have declared no competing interest.

